# Factors associated with health-related quality of life in people with severe mental illness: Results from a multicountry study in South Asia

**DOI:** 10.1101/2025.08.01.25332563

**Authors:** Avantika Sharma, Badur Un Nisa, Humaira Bibi, Fraser Wiggins, Alex Mitchell, Krishna Prasad, Kavindu Appuhamy, Olga P Garcia, Gerardo A Zavala

## Abstract

**Background:** Health-related quality of life (HRQoL) is a multidimensional measure of well-being encompassing physical, psychological, and social domains. Individuals with severe mental illness (SMI) experience substantial HRQoL challenges, particularly in low- and middle-income countries (LMICs) where healthcare disparities persist. This study examines sociodemographic, economic, and health-related factors associated with HRQoL in people with SMI across Bangladesh, India, and Pakistan.

**Methods:** A cross-sectional study was conducted as part of the IMPACT survey, recruiting 3,989 participants with SMI. HRQoL was assessed using the EQ-5D-5L tool, covering mobility, self-care, usual activities, pain/discomfort, and anxiety/depression. Logistic regression models explored associations between HRQoL and demographic, socioeconomic, and health factors, including BMI and smoking status.

**Results:** HRQoL varied significantly by sociodemographic and geographic factors. Participants from India had lower odds of HRQoL difficulties than those in Pakistan and Bangladesh. Higher income and education were linked to better HRQoL, while female gender, underweight status, unemployment, and smoking were associated with significantly worse outcomes. For instance, underweight individuals had 1.30 times higher odds (95% CI=0.98- 1.73, p=0.072) of mobility problems, while smokers had 1.34 times higher odds (95% CI=1.13- 1.59, p<0.001) of self-care difficulties. Women reported worse HRQoL than men.

**Conclusion:** Addressing income disparities, promoting education, supporting employment, and reducing smoking prevalence is essential to improve HRQoL in people with SMI in South Asia. Targeted interventions that consider geographical and demographic contexts can enhance quality of life in people attending mental health facilities in LMICs.

## Introduction

Health-related quality of life (HRQoL) is an essential construct in the care and evaluation of individuals with severe mental illness (SMI), encompassing physical, psychological, social, and functional well-being [1]. Individuals with SMI frequently experience persistent symptoms, social stigma, and physical health complications related to both their condition and its treatment. Understanding HRQoL in this population provides critical insight into overall health status and day-to-day functioning [2]. Improved HRQoL reflects better symptom management and the ability to participate in meaningful activities, whereas diminished HRQoL may indicate unmet clinical or social needs.

Recent literature has increasingly examined factors associated with HRQoL in people with SMI [3–5]. However, evidence from low- and middle-income countries (LMICs) remains limited. Contextual factors in LMICs, such as healthcare disparities, sociocultural norms, and resource constraints, may influence these associations differently than in high-income settings. Notably, the relationship between obesity and HRQoL in individuals with SMI has not been investigated in South Asia, despite the unique cultural, economic, and healthcare landscapes that could significantly shape outcomes [6–8].

Although the burden of SMI in LMICs is substantial, healthcare systems often focus narrowly on psychiatric symptom management, neglecting physical health needs [9,10]. Individuals with SMI in LMICs encounter numerous barriers, including inadequate access to healthcare, poverty, and stigma, all of which may adversely affect physical health and HRQoL [11–13]. Despite the known intersection of physical illness and mental health, little research has explored how health-risk behaviours and comorbid physical conditions influence HRQoL among people with SMI in these settings [14].

Previous research by Niazi et al. (2023) analysed data from Pakistan using the EQ-5D visual analogue scale (VAS) to explore HRQoL and described domain-level outcomes in relation to SMI conditions. While their study offered important insights, it was limited to a single-country analysis and focused primarily on descriptive associations using the VAS [15]. Building on this work, the present study expands the scope by utilising the full dataset from India, Pakistan, and Bangladesh.

Beyond descriptive analyses, this study examines associations between sociodemographic and clinical factors and each EQ-5D HRQoL domain. By incorporating data from India, Pakistan, and Bangladesh, it offers a broader regional perspective and deeper analytic insight into HRQoL among individuals with SMI in South Asia. Addressing this gap is essential for developing context-specific strategies to improve health outcomes and reduce disparities. A clearer understanding of these determinants can guide clinicians and policymakers, support targeted interventions, and enhance patient-centred care. Specifically, this study investigates the prevalence of HRQoL impairments and their associations with demographic, socioeconomic, and health-related factors, including obesity, in people with SMI across the region.

## Methods

### Study Design

We conducted a cross-sectional analysis using baseline data from the IMPACT study (ISRCTN registry: 88485933), a multi-country survey examining health, health risk behaviours, and healthcare utilisation among individuals with SMI in India, Bangladesh, and Pakistan. Further details of the methods are reported in the published protocol [16].

### Study Setting

Participants were recruited from three tertiary mental health institutions: the National Institute of Mental Health and Neurosciences (NIMHANS) in Bengaluru, India; the National Institute of Mental Health and Hospital (NIMHH) in Dhaka, Bangladesh; and the Institute of Psychiatry (IOP) at Rawalpindi Medical University in Pakistan. These facilities, while classified as tertiary centres, provide services to the general population due to limited mental health services at lower levels of care [17].

### Eligibility Criteria

Participants were adults (aged ≥18 years) diagnosed with SMI by local psychiatrists and capable of providing informed consent. Diagnoses were based on the International Classification of Diseases, 10th Revision (ICD-10) codes F20–F29, F30–F31, and F32.3– F33.3 [18]. Diagnoses were validated using the Mini-International Neuropsychiatric Interview (MINI) version 6.0, a structured diagnostic tool suitable for use by trained non-specialists [19,20].

### Sampling and Recruitment

Stratified random sampling was used to select participants from outpatient and inpatient services. Random number tables were centrally generated to determine which individuals or beds to sample [19,20]. The targeted recruitment ratio was 80% outpatients and 20% inpatients, reflecting the typical distribution of care settings for individuals with SMI. Reimbursement for their travel expenses and the results from the clinical investigation were given.

### Data Collection

Trained mental health researchers conducted face-to-face digital interviews in local languages (Urdu, Bangla, Hindi, Kannada, Tamil, and Telugu). The World Health Organisation STEPwise approach to Surveillance (WHO STEPs) was used to collect sociodemographic data [23]. Interviews were conducted by researchers fluent in local languages, and participants could request interviewers of the same gender to reduce potential response bias [17]. Physical measurements, including height, weight, and waist circumference, were recorded using standardised protocols.

### Data Availability

The data supporting the findings of this study are available upon reasonable request from the corresponding author. Due to ethical restrictions, data cannot be made publicly available.

### Patient and Public Involvement

Patients, caregivers, and advocacy group members formed a community advisory panel that reviewed and piloted the survey instrument, ensuring its cultural and contextual relevance.

### Sample Size Calculation

Sample size calculations were initially based on estimating a diabetes prevalence of 10% with 2% precision at the 95% confidence level, yielding a target of 857 participants per country [16,21,22]. This sample was also sufficient for estimating HRQoL outcome with <2% margin of error.

### Ethical Considerations

Ethical approvals were obtained from the relevant institutional review boards, including the University of York, the Indian Council of Medical Research, Bangladesh’s Centre for Injury Prevention and Rehabilitation, and Pakistan’s National Bioethics Committee [16]. Informed consent was obtained from all participants.

### Dependent Variable: HRQoL

HRQoL was assessed using the EQ-5D-5L instrument, comprising five domains: mobility, self-care, usual activities, pain/discomfort, and anxiety/depression. Each domain includes five response levels. For analysis, responses were dichotomised into "no problems" and "slight to severe problems." The instrument also includes a visual analogue scale (VAS) ranging from 0 (worst imaginable health) to 100 (best imaginable health) [24].

### Independent Variables

Independent variables included BMI (calculated using both WHO international and Asian-specific cut-offs [25–27]), age, sex, education, employment status, income, diagnosis and duration of SMI, comorbid physical conditions (e.g., diabetes, hypertension), antipsychotic medication use, clinical setting (inpatient vs outpatient), and smoking status.

### Statistical Analysis

Descriptive statistics summarise sample characteristics. Categorical variables were presented as frequencies and percentages; continuous variables as means and standard deviations. Prevalence of HRQoL impairments was stratified by country and BMI category. Multivariate logistic regression models examined associations between BMI and each HRQoL domain, adjusting for demographic and health-related covariates. Odds ratios (ORs) and 95% confidence intervals (CIs) were reported. Analyses were conducted using Stata version 17.0, with significance set at p < 0.05.

## Results

Table 1 presents the sociodemographic, clinical, and anthropometric characteristics of the study sample (N = 3,989), stratified by country. The sample was predominantly male (59.1%), with a mean age of 35.8 years (SD = 11.9). The largest age group was 25–39 years (47.0%), followed by those aged 40–54 years (26.3%). Educational attainment was low overall, with 13.8% of participants reporting no formal education, and only 30.7% completing education beyond the secondary level.

**Table 1:**
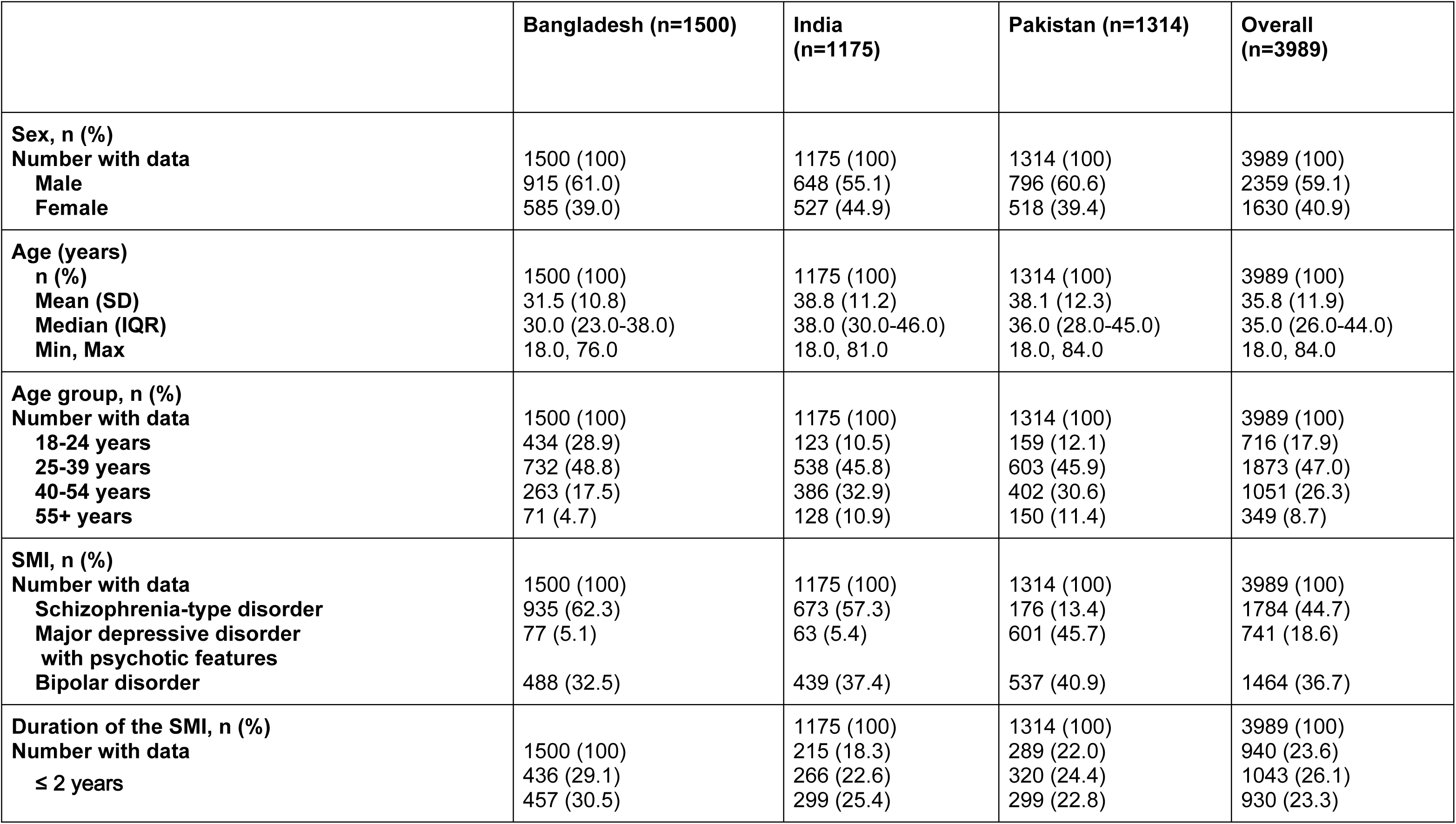
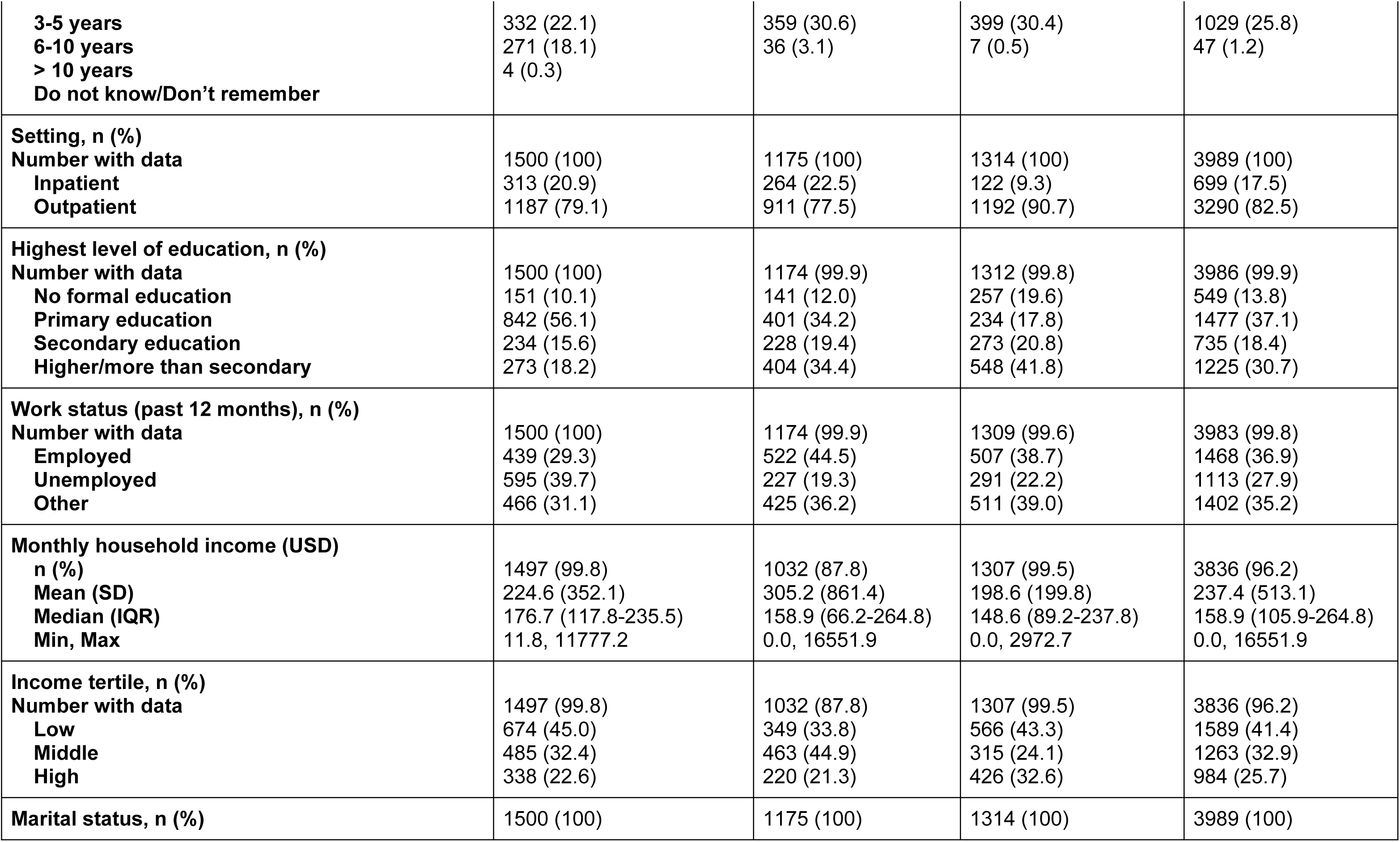
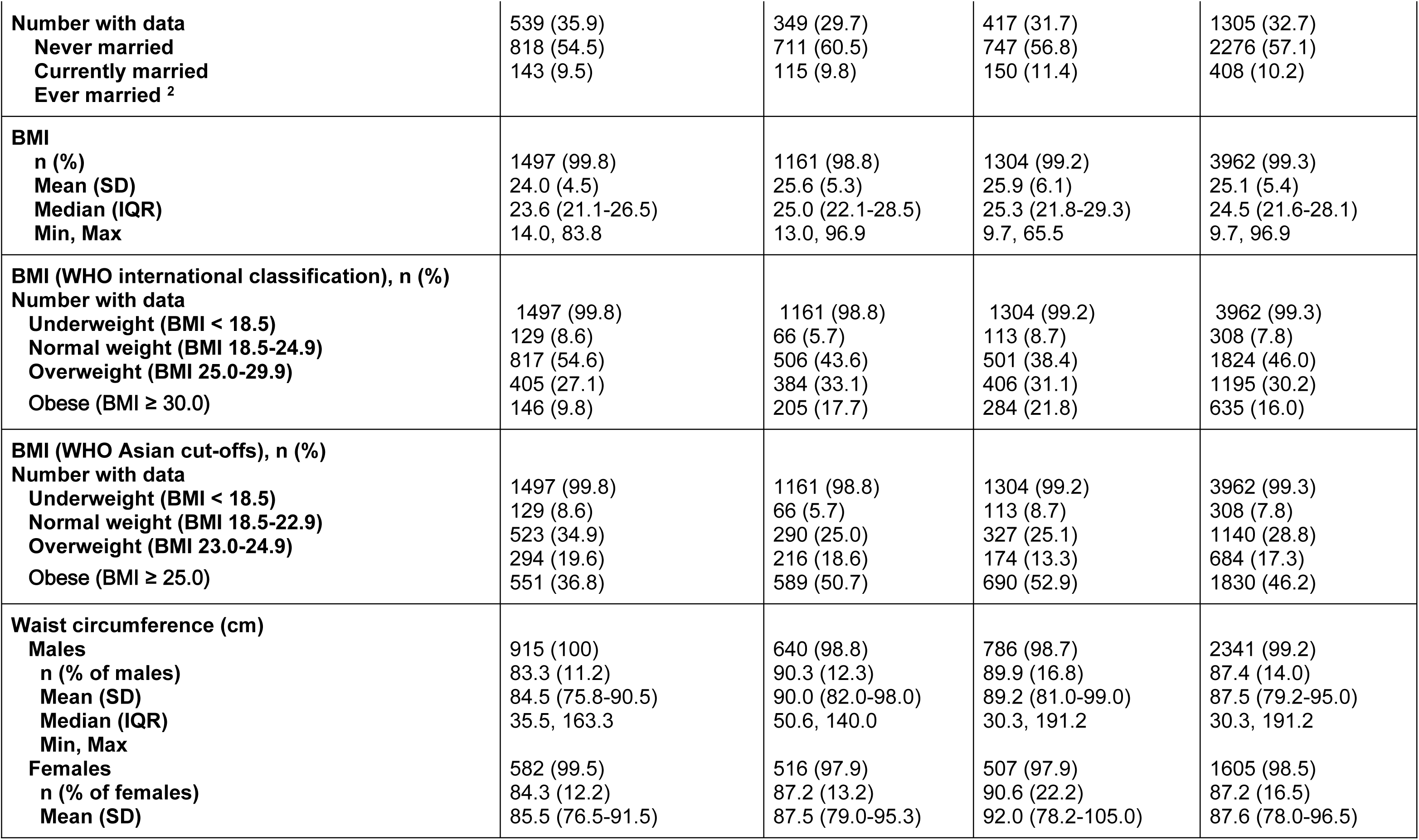
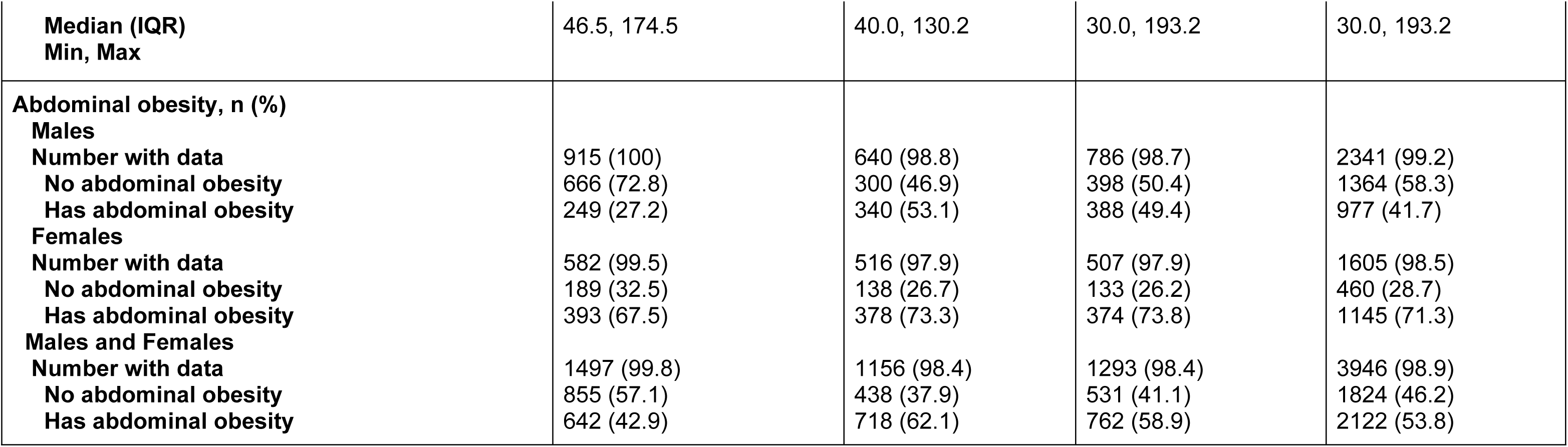
Participant characteristics summarised by country.

Most participants were diagnosed with schizophrenia-spectrum disorders (44.7%), followed by bipolar disorder (36.7%) and major depressive disorder with psychotic features (18.6%). Approximately 25.8% of the sample had been living with SMI for over 10 years. The majority of participants were recruited from outpatient settings (82.5%).

Regarding socioeconomic status, 41.4% of participants belonged to the lowest income tertile, and the mean monthly household income was USD 237.4 (SD = 513.1). Employment rates were suboptimal: only 36.9% were employed, while 27.9% were unemployed and 35.2% fell into the “other” category (including students, homemakers, and retirees). Marital status varied across the sample, with 57.1% currently married and 32.7% never married.

From a physical health perspective, the mean BMI was 25.1 kg/m² (SD = 5.4), but the distribution differed depending on the cut-offs used. Based on WHO international standards, 16.0% of participants were classified as obese and 7.8% as underweight. Using WHO Asian cut-offs, the obesity prevalence rose to 46.2%, underscoring the higher cardiometabolic risk profile in South Asian populations. Smoking was highly prevalent (34.5%), particularly among men (52.3%), while alcohol use was minimal (6.8%), likely reflecting cultural and religious norms. Chronic physical conditions were common: 23.2% of participants had hypertension, 18.5% had diabetes, and 12.7% had dyslipidemia.

### Prevalence of HRQoL Problems

The data reveal consistent patterns of inequality in health-related quality of life (HRQoL) across sociodemographic and health-status indicators (Table 2).

**Table 2:** Health-Related Quality of Life Domain Problem vs No Problem.

|  | Mobility |  | Self-Care |  | Usual Activities |  | Pain/discomfort |  | Anxiety/depression |  |
| --- | --- | --- | --- | --- | --- | --- | --- | --- | --- | --- |
|  | No problem | Problem | No problem | Problem | No problem | Problem | No problem | Problem | No problem | Problem |
| **WHO International BMI Cut-offs, n (%)** |  |  |  |  |  |  |  |  |  |  |
| **Underweight (BMI < 18.5)** | 176/308<br>(57.1) | 132/308<br>(42.9) | 168/308<br>(54.5) | 140/308<br>(45.5) | 139/308<br>(45.1) | 169/308<br>(54.9) | 100/308<br>(32.5) | 208/308<br>(67.5) | 73/308<br>(23.7) | 235/308<br>(76.3) |
| **Normal weight (BMI 18.5-24.9)** | 1204/1824<br>(66.0) | 620/1824<br>(34.0) | 1175/1824<br>(64.4) | 649/1824<br>(35.6) | 997/1824<br>(54.7) | 827/1824<br>(45.3) | 732/1824<br>(40.1) | 1092/1824<br>(59.9) | 508/1824<br>(27.9) | 1316/1824<br>(72.1) |
| **Overweight (BMI 25.0-29.9)** | 787/1195<br>(65.9) | 408/1195<br>(34.1) | 829/1195<br>(69.4) | 366/1195<br>(30.6) | 702/1195<br>(58.7) | 493/1195<br>(41.3) | 467/1195<br>(39.1) | 728/1195<br>(60.9) | 351/1195<br>(29.4) | 844/1195<br>(70.6) |
| **Obese (BMI ≥30.0)** | 376/635<br>(59.2) | 259/635<br>(40.8) | 428/635<br>(67.4) | 207/635<br>(32.6) | 366/635<br>(57.6) | 269/635<br>(42.4) | 238/635<br>(37.5) | 397/635<br>(62.5) | 216/635<br>(34.0) | 419/635<br>(66.0) |
| **Education Status, n (%)** |  |  |  |  |  |  |  |  |  |  |
| **No formal education** | 284/549<br>(51.7) | 265/549<br>(48.3) | 301/549<br>(54.8) | 248/549<br>(45.2) | 248/549<br>(45.2) | 301/549<br>(54.8) | 159/549<br>(29.0) | 390/549<br>(71.0) | 149/549<br>(27.1) | 400/549<br>(72.9) |
| **Primary education** | 975/1477<br>(66.0) | 502/1477<br>(34.0) | 987/1477<br>(66.8) | 490/1477<br>(33.2) | 846/1477<br>(57.3) | 631/1477<br>(42.7) | 579/1477<br>(39.2) | 898/1477<br>(60.8) | 388/1477<br>(26.3) | 1089/1477<br>(73.7) |
| **Secondary education** | 484/735<br>(65.9) | 251/735<br>(34.1) | 508/735<br>(69.1) | 227/735<br>(30.9) | 433/735<br>(58.9) | 302/735<br>(41.1) | 310/735<br>(42.2) | 425/735<br>(57.8) | 227/735<br>(30.9) | 508/735<br>(69.1) |
| **Higher/more than secondary education** | 812/1225<br>(66.3) | 413/1225<br>(33.7) | 817/1225<br>(66.7) | 408/1225<br>(33.3) | 686/1225<br>(56.0) | 539/1225<br>(44.0) | 498/1225<br>(40.7) | 727/1225<br>(59.3) | 391/1225<br>(31.9) | 834/1225<br>(68.1) |
| **Employment Status, n (%)** |  |  |  |  |  |  |  |  |  |  |
| **Unemployed** | 693/1113<br>(62.3) | 420/1113<br>(37.7) | 624/1113<br>(56.1) | 489/1113<br>(43.9) | 469/1113<br>(42.1) | 644/1113<br>(57.9) | 364/1113<br>(32.7) | 749/1113<br>(67.3) | 213/1113<br>(19.1) | 900/1113<br>(80.9) |
| **Employed** | 1059/1468<br>(72.1) | 409/1468<br>(27.9) | 1116/1468<br>(76.0) | 352/1468<br>(24.0) | 998/1468<br>(68.0) | 470/1468<br>(32.0) | 680/1468<br>(46.3) | 788/1468<br>(53.7) | 542/1468<br>(36.9) | 926/1468<br>(63.1) |
| **Other (student, homemaker, retired)** | 803/1402<br>(57.3) | 599/1402<br>(42.7) | 873/1402<br>(62.3) | 529/1402<br>(37.7) | 744/1402<br>(53.1) | 658/1402<br>(46.9) | 503/1402<br>(35.9) | 899/1402<br>(64.1) | 401/1402<br>(28.6) | 1001/1402<br>(71.4) |
| **Income Tertile, n (%)** |  |  |  |  |  |  |  |  |  |  |
| **Low** | 992/1589<br>(62.4) | 597/1589<br>(37.6) | 1020/1589<br>(64.2) | 569/1589<br>(35.8) | 825/1589<br>(51.9) | 764/1589<br>(48.1) | 513/1589<br>(32.3) | 1076/1589<br>(67.7) | 366/1589<br>(23.0) | 1223/1589<br>(77.0) |
| **Middle** | 814/1263<br>(64.4) | 449/1263<br>(35.6) | 832/1263<br>(65.9) | 431/1263<br>(34.1) | 703/1263<br>(55.7) | 560/1263<br>(44.3) | 530/1263<br>(42.0) | 733/1263<br>(58.0) | 397/1263<br>(31.4) | 866/1263<br>(68.6) |
| **High** | 639/984<br>(64.9) | 345/984<br>(35.1) | 639/984<br>(64.9) | 345/984<br>(35.1) | 571/984<br>(58.0) | 413/984<br>(42.0) | 402/984<br>(40.9) | 582/984<br>(59.1) | 297/984<br>(30.2) | 687/984<br>(69.8) |

Participants with underweight BMI (BMI < 18.5) reported the highest proportion of impairments across nearly all domains, particularly in self-care (45.5%) and usual activities (54.9%), highlighting the functional limitations associated with poor nutritional and physical health. In contrast, those in the overweight and obese categories showed somewhat lower levels of self-care problems (30.6% and 32.6%, respectively), supporting findings from Table 3 that suggest a potential “protective” effect of overweight status on certain functional domains.

**Table 3:** Logistic Regression Analysis of Health-Related Quality of Life (domain) Issues Related to Participant Characteristics.

|  | Mobility |  | Self-Care |  | Usual Activities |  | Pain/discomfort |  | Anxiety/depression |  |
| --- | --- | --- | --- | --- | --- | --- | --- | --- | --- | --- |
|  | Frequency with problems | OR (95% CI), p-value | Frequency with problems | OR (95% CI), p-value | Frequency with problems | OR (95% CI), p-value | Frequency with problems | OR (95% CI), p-value | Frequency with problems | OR (95% CI), p-value |
| **WHO International BMI Cut-offs** |  |  |  |  |  |  |  |  |  |  |
| **Underweight (BMI < 18.5)** | 118/273 (43.2) | 1.30 (0.98-1.73), p=0.072 | 124/273 (45.4) | 1.24 (0.94-1.64), p=0.128 | 152/273 (55.7) | 1.24 (0.94-1.64), p=0.121 | 188/273 (68.9) | 1.22 (0.91-1.64), p=0.192 | 212/273 (77.7) | 0.94 (0.66-1.34), p=0.741 |
| **Normal weight (BMI 18.5-24.9)** | 552/1583 (34.9) | Reference | 580/1583 (36.6) | Reference | 737/1583 (46.6) | Reference | 982/1583 (62.0) | Reference | 1198/1583 (75.7) | Reference |
| **Overweight (BMI 25.0-29.9)** | 355/1007 (35.3) | 0.94 (0.78-1.13), p=0.512 | 311/1007 (30.9) | 0.78 (0.65-0.94), p=0.009 | 428/1007 (42.5) | 0.88 (0.74-1.04), p=0.142 | 642/1007 (63.8) | 1.10 (0.91-1.32), p=0.314 | 752/1007 (74.7) | 1.15 (0.93-1.43), p=0.207 |
| **Obese (BMI ≥30.0)** | 229/536 (42.7) | 1.07 (0.85-1.36), p=0.547 | 186/536 (34.7) | 0.81 (0.64-1.03), p=0.085 | 246/536 (45.9) | 0.89 (0.71-1.11), p=0.310 | 357/536 (66.6) | 1.11 (0.87-1.42), p=0.384 | 365/536 (68.1) | 0.83 (0.63-1.08), p=0.162 |
| **Sex** |  |  |  |  |  |  |  |  |  |  |
| **Male** | 671/2034 (33.0) | Reference | 676/2034 (33.2) | Reference | 887/2034 (43.6) | Reference | 1236/2034 (60.8) | Reference | 1476/2034 (72.6) | Reference |
| **Female** | 583/1365 (42.7) | 1.08 (0.85-1.37), p=0.525 | 525/1365 (38.5) | 1.08 (0.85-1.37), p=0.520 | 676/1365 (49.5) | 1.15 (0.92-1.44), p=0.234 | 933/1365 (68.4) | 1.51 (1.19-1.91), p<0.001 | 1051/1365 (77.0) | 1.66 (1.24-2.23), p<0.001 |
| **Age Group** |  |  |  |  |  |  |  |  |  |  |
| **18-24 years** | 197/642 (30.7) | Reference | 230/642 (35.8) | Reference | 290/642 (45.2) | Reference | 393/642 (61.2) | Reference | 520/642 (81.0) | Reference |
| **25-39 years** | 545/1600 (34.1) | 1.34 (1.07-1.67), p=0.011 | 525/1600 (32.8) | 0.98 (0.79-1.22), p=0.889 | 716/1600 (44.8) | 1.12 (0.91-1.38), p=0.289 | 983/1600 (61.4) | 1.18 (0.95-1.47), p=0.127 | 1191/1600 (74.4) | 1.09 (0.83-1.45), p=0.536 |
| **40-59 years** | 354/867 (40.8) | 1.64 (1.26-2.14), p<0.001 | 311/867 (35.9) | 1.10 (0.85-1.43), p=0.481 | 402/867 (46.4) | 1.17 (0.91-1.50), p=0.226 | 581/867 (67.0) | 1.55 (1.19-2.03), p=0.001 | 621/867 (71.6) | 1.23 (0.89-1.70), p=0.209 |
| **55+ years** | 158/290<br>(54.5) | 2.70 (1.89-3.85),<br>p<0.001 | 135/290<br>(46.6) | 1.49 (1.05-2.11),<br>p=0.026 | 155/290<br>(53.4) | 1.33 (0.94-1.87),<br>p=0.103 | 212/290<br>(73.1) | 2.10 (1.44-3.07),<br>p<0.001 | 195/290<br>(67.2) | 0.95 (0.62-1.44),<br>p=0.802 |
| **SMI Diagnosis** |  |  |  |  |  |  |  |  |  |  |
| **Bipolar disorder (any)** | 391/1245<br>(31.4) | Reference | 343/1245<br>(27.6) | Reference | 470/1245<br>(37.8) | Reference | 756/1245<br>(60.7) | Reference | 837/1245<br>(67.2) | Reference |
| **Major depressive disorder with psychotic features** | 417/670<br>(62.2) | 2.34 (1.87-2.92),<br>p<0.001 | 344/670<br>(51.3) | 1.99 (1.59-2.49),<br>p<0.001 | 397/670<br>(59.3) | 1.79 (1.44-2.24),<br>p<0.001 | 559/670<br>(83.4) | 2.24 (1.73-2.91),<br>p<0.001 | 597/670<br>(89.1) | 5.00 (3.71-6.75),<br>p<0.001 |
| **Psychotic disorder** | 446/1484<br>(30.1) | 1.21 (1.01-1.46),<br>p=0.044 | 514/1484<br>(34.6) | 1.65 (1.37-2.00),<br>p<0.001 | 696/1484<br>(46.9) | 1.60 (1.34-1.91),<br>p<0.001 | 854/1484<br>(57.5) | 0.96 (0.81-1.15),<br>p=0.658 | 1093/1484<br>(73.7) | 0.95 (0.77-1.17),<br>p=0.630 |
| **Setting** |  |  |  |  |  |  |  |  |  |  |
| **Inpatient** | 217/529<br>(41.0) | Reference | 228/529<br>(43.1) | Reference | 304/529<br>(57.5) | Reference | 388/529<br>(73.3) | Reference | 447/529<br>(84.5) | Reference |
| **Outpatient** | 1037/2870<br>(36.1) | 0.58 (0.47-0.72),<br>p<0.001 | 973/2870<br>(33.9) | 0.55 (0.45-0.68),<br>p<0.001 | 1259/2870<br>(43.9) | 0.52 (0.42-0.63),<br>p<0.001 | 1781/2870<br>(62.1) | 0.43 (0.34-0.54),<br>p<0.001 | 2080/2870<br>(72.5) | 0.42 (0.31-0.56),<br>p<0.001 |
| **Level of Education** |  |  |  |  |  |  |  |  |  |  |
| **No formal education** | 237/474<br>(50.0) | Reference | 226/474<br>(47.7) | Reference | 275/474<br>(58.0) | Reference | 354/474<br>(74.7) | Reference | 362/474<br>(76.4) | Reference |
| **Primary education** | 441/1271<br>(34.7) | 0.98 (0.76-1.26),<br>p=0.862 | 425/1271<br>(33.4) | 0.76 (0.59-0.97),<br>p=0.027 | 554/1271<br>(43.6) | 0.70 (0.55-0.90),<br>p=0.004 | 799/1271<br>(62.9) | 0.87 (0.66-1.13),<br>p=0.296 | 986/1271<br>(77.6) | 0.92 (0.68-1.24),<br>p=0.568 |
| **Secondary education** | 211/624<br>(33.8) | 0.76 (0.58-1.01),<br>p=0.061 | 196/624<br>(31.4) | 0.64 (0.48-0.84),<br>p=0.002 | 263/624<br>(42.1) | 0.65 (0.50-0.85),<br>p=0.002 | 371/624<br>(59.5) | 0.72 (0.54-0.97),<br>p=0.032 | 447/624<br>(71.6) | 1.03 (0.74-1.43),<br>p=0.863 |
| **Higher/more than secondary education** | 365/1030<br>(35.4) | 0.74 (0.57-0.97),<br>p=0.027 | 354/1030<br>(34.4) | 0.71 (0.55-0.91),<br>p=0.008 | 471/1030<br>(45.7) | 0.75 (0.58-0.97),<br>p=0.026 | 645/1030<br>(62.6) | 0.84 (0.63-1.12),<br>p=0.235 | 732/1030<br>(71.1) | 1.07 (0.78-1.46),<br>p=0.663 |
| **Work status (past 12 months)** |  |  |  |  |  |  |  |  |  |  |
| **Employed** | 356/1247<br>(28.5) | Reference | 305/1247<br>(24.5) | Reference | 416/1247<br>(33.4) | Reference | 706/1247<br>(56.6) | Reference | 832/1247<br>(66.7) | Reference |
| **Unemployed** | 367/967<br>(38.0) | 1.67 (1.37-2.05),<br>p<0.001 | 428/967<br>(44.3) | 2.30 (1.88-2.81),<br>p<0.001 | 563/967<br>(58.2) | 2.58 (2.13-3.13),<br>p<0.001 | 660/967<br>(68.3) | 1.43 (1.17-1.75),<br>p<0.001 | 802/967<br>(82.9) | 1.61 (1.27-2.05),<br>p<0.001 |
| **Other (student, homemaker, retired)** | 531/1185<br>(44.8) | 1.76 (1.35-2.28),<br>p<0.001 | 468/1185<br>(39.5) | 1.82 (1.40-2.36),<br>p<0.001 | 584/1185<br>(49.3) | 1.71 (1.34-2.19),<br>p<0.001 | 803/1185<br>(67.8) | 1.11 (0.86-1.43),<br>p=0.429 | 893/1185<br>(75.4) | 0.93 (0.69-1.26),<br>p=0.648 |
| **Income tertile** |  |  |  |  |  |  |  |  |  |  |
| **Low** | 534/1442<br>(37.0) | Reference | 513/1442<br>(35.6) | Reference | 692/1442<br>(48.0) | Reference | 983/1442<br>(68.2) | Reference | 1124/1442<br>(77.9) | Reference |
| **Middle** | 405/1080<br>(37.5) | 1.27 (1.06-1.52),<br>p=0.009 | 374/1080<br>(34.6) | 1.21 (1.01-1.45),<br>p=0.038 | 499/1080<br>(46.2) | 1.13 (0.96-1.35),<br>p=0.148 | 658/1080<br>(60.9) | 0.91 (0.76-1.09),<br>p=0.309 | 774/1080<br>(71.7) | 0.88 (0.71-1.10),<br>p=0.261 |
| **High** | 315/877<br>(35.9) | 0.87 (0.71-1.06),<br>p=0.172 | 314/877<br>(35.8) | 1.04 (0.85-1.26),<br>p=0.710 | 372/877<br>(42.4) | 0.81 (0.67-0.97),<br>p=0.024 | 528/877<br>(60.2) | 0.65 (0.53-0.79),<br>p<0.001 | 629/877<br>(71.7) | 0.70 (0.56-0.88),<br>p=0.003 |
| **Country** |  |  |  |  |  |  |  |  |  |  |
| **Bangladesh** | 439/1421<br>(30.9) | Reference | 495/1421<br>(34.8) | Reference | 639/1421<br>(45.0) | Reference | 934/1421<br>(65.7) | Reference | 1298/1421<br>(91.3) | Reference |
| **India** | 151/749<br>(20.2) | 0.58 (0.46-0.73),<br>p<0.001 | 134/749<br>(17.9) | 0.46 (0.37-0.59),<br>p<0.001 | 246/749<br>(32.8) | 0.67 (0.54-0.83),<br>p<0.001 | 290/749<br>(38.7) | 0.30 (0.24-0.37),<br>p<0.001 | 333/749<br>(44.5) | 0.07 (0.05-0.09),<br>p<0.001 |
| **Pakistan** | 664/1229<br>(54.0) | 2.47 (1.98-3.08),<br>p<0.001 | 572/1229<br>(46.5) | 1.93 (1.55-2.41),<br>p<0.001 | 678/1229<br>(55.2) | 1.76 (1.42-2.18),<br>p<0.001 | 945/1229<br>(76.9) | 1.40 (1.12-1.75),<br>p=0.003 | 896/1229<br>(72.9) | 0.15 (0.11-0.19),<br>p<0.001 |
| **Five fruit and veg consumed per day** |  |  |  |  |  |  |  |  |  |  |
| **No** | 1196/3224<br>(37.1) | Reference | 1138/3224<br>(35.3) | Reference | 1480/3224<br>(45.9) | Reference | 2066/3224<br>(64.1) | Reference | 2389/3224<br>(74.1) | Reference |
| **Yes** | 58/175<br>(33.1) | 0.93 (0.66-1.32),<br>p=0.685 | 63/175<br>(36.0) | 1.08 (0.77-1.51),<br>p=0.669 | 83/175<br>(47.4) | 1.16 (0.83-1.61),<br>p=0.378 | 103/175<br>(58.9) | 0.76 (0.54-1.06),<br>p=0.104 | 138/175<br>(78.9) | 0.86 (0.56-1.31),<br>p=0.479 |
| **High blood pressure** |  |  |  |  |  |  |  |  |  |  |
| **No** | 991/2871<br>(34.5) | Reference | 987/2871<br>(34.4) | Reference | 1290/2871<br>(44.9) | Reference | 1783/2871<br>(62.1) | Reference | 2126/2871<br>(74.1) | Reference |
| **Yes** | 263/528<br>(49.8) | 1.35 (1.09-1.68),<br>p=0.006 | 214/528<br>(40.5) | 1.06 (0.85-1.32),<br>p=0.608 | 273/528<br>(51.7) | 1.18 (0.95-1.45),<br>p=0.128 | 386/528<br>(73.1) | 1.30 (1.03-1.65),<br>p=0.028 | 401/528<br>(75.9) | 1.30 (1.01-1.69),<br>p=0.042 |
| **High blood sugar** |  |  |  |  |  |  |  |  |  |  |
| **No** | 1107/3056<br>(36.2) | Reference | 1063/3056<br>(34.8) | Reference | 1388/3056<br>(45.4) | Reference | 1942/3056<br>(63.5) | Reference | 2276/3056<br>(74.5) | Reference |
| **Yes** | 147/343<br>(42.9) | 1.14 (0.87-1.49),<br>p=0.345 | 138/343<br>(40.2) | 1.27 (0.97-1.65),<br>p=0.079 | 175/343<br>(51.0) | 1.20 (0.93-1.54),<br>p=0.170 | 227/343<br>(66.2) | 1.08 (0.82-1.43),<br>p=0.589 | 251/343<br>(73.2) | 1.06 (0.77-1.45),<br>p=0.732 |
| **High Cholesterol** |  |  |  |  |  |  |  |  |  |  |
| **No** | 603/1686<br>(35.8) | Reference | 591/1686<br>(35.1) | Reference | 804/1686<br>(47.7) | Reference | 1073/1686<br>(63.6) | Reference | 1270/1686<br>(75.3) | Reference |
| **Yes** | 651/1713<br>(38.0) | 0.94 (0.81-1.10),<br>p=0.454 | 610/1713<br>(35.6) | 0.98 (0.84-1.14),<br>p=0.761 | 759/1713<br>(44.3) | 0.82 (0.71-0.95),<br>p=0.008 | 1096/1713<br>(64.0) | 0.91 (0.78-1.07),<br>p=0.260 | 1257/1713<br>(73.4) | 0.90 (0.75-1.08),<br>p=0.245 |
| **High Triglycerides** |  |  |  |  |  |  |  |  |  |  |
| **No** | 744/2107<br>(35.3) | Reference | 726/2107<br>(34.5) | Reference | 936/2107<br>(44.4) | Reference | 1299/2107<br>(61.7) | Reference | 1538/2107<br>(73.0) | Reference |
| **Yes** | 510/1292<br>(39.5) | 1.15 (0.98-1.35),<br>p=0.080 | 475/1292<br>(36.8) | 1.08 (0.92-1.26),<br>p=0.343 | 627/1292<br>(48.5) | 1.19 (1.02-1.38),<br>p=0.027 | 870/1292<br>(67.3) | 1.20 (1.02-1.41),<br>p=0.029 | 989/1292<br>(76.5) | 1.08 (0.89-1.31),<br>p=0.420 |
| **Meets WHO physical activity recommendations** |  |  |  |  |  |  |  |  |  |  |
| **No** | 667/1767<br>(37.7) | Reference | 644/1767<br>(36.4) | Reference | 872/1767<br>(49.3) | Reference | 1134/1767<br>(64.2) | Reference | 1290/1767<br>(73.0) | Reference |
| **Yes** | 587/1632<br>(36.0) | 1.23 (1.04-1.44),<br>p=0.013 | 557/1632<br>(34.1) | 1.05 (0.89-1.23),<br>p=0.563 | 691/1632<br>(42.3) | 0.85 (0.73-0.99),<br>p=0.032 | 1035/1632<br>(63.4) | 1.03 (0.88-1.21),<br>p=0.704 | 1237/1632<br>(75.8) | 0.84 (0.70-1.02),<br>p=0.076 |
| **Indication of Pre-diabetes** |  |  |  |  |  |  |  |  |  |  |
| **No** | 979/2637<br>(37.1) | Reference | 940/2637<br>(35.6) | Reference | 1232/2637<br>(46.7) | Reference | 1678/2637<br>(63.6) | Reference | 1964/2637<br>(74.5) | Reference |
| **Yes** | 275/762<br>(36.1) | 1.06 (0.88-1.28),<br>p=0.539 | 261/762<br>(34.3) | 1.04 (0.86-1.25),<br>p=0.694 | 331/762<br>(43.4) | 0.93 (0.77-1.11),<br>p=0.395 | 491/762<br>(64.4) | 1.11 (0.91-1.34),<br>p=0.301 | 563/762<br>(73.9) | 0.90 (0.72-1.12),<br>p=0.354 |
| **Received Medication for SMI** |  |  |  |  |  |  |  |  |  |  |
| **No** | 46/104<br>(44.2) | Reference | 42/104<br>(40.4) | Reference | 54/104<br>(51.9) | Reference | 67/104<br>(64.4) | Reference | 82/104<br>(78.8) | Reference |
| **Yes** | 1208/3295<br>(36.7) | 0.98 (0.63-1.51),<br>p=0.914 | 1159/3295<br>(35.2) | 1.04 (0.67-1.60),<br>p=0.866 | 1509/3295<br>(45.8) | 0.91 (0.60-1.40),<br>p=0.678 | 2102/3295<br>(63.8) | 1.26 (0.80-1.97),<br>p=0.316 | 2445/3295<br>(74.2) | 0.82 (0.48-1.40),<br>p=0.469 |
| **Smoking Status** |  |  |  |  |  |  |  |  |  |  |
| **No** | 802/2153<br>(37.3) | Reference | 716/2153<br>(33.3) | Reference | 946/2153<br>(43.9) | Reference | 1339/2153<br>(62.2) | Reference | 1573/2153<br>(73.1) | Reference |
| **Yes** | 452/1246<br>(36.3) | 0.96 (0.81-1.14),<br>p=0.653 | 485/1246<br>(38.9) | 1.34 (1.13-1.59),<br>p<0.001 | 617/1246<br>(49.5) | 1.35 (1.14-1.59),<br>p<0.001 | 830/1246<br>(66.6) | 1.18 (0.99-1.40),<br>p=0.066 | 954/1246<br>(76.6) | 1.17 (0.95-1.44),<br>p=0.134 |
| **Duration of SMI in years** |  |  |  |  |  |  |  |  |  |  |
| **<= 2 years** | 319/802<br>(39.8) | Reference | 303/802<br>(37.8) | Reference | 382/802<br>(47.6) | Reference | 534/802<br>(66.6) | Reference | 647/802<br>(80.7) | Reference |
| **3-5 years** | 340/910<br>(37.4) | 0.97 (0.78-1.20),<br>p=0.756 | 321/910<br>(35.3) | 0.96 (0.78-1.19),<br>p=0.724 | 415/910<br>(45.6) | 0.96 (0.78-1.18),<br>p=0.705 | 583/910<br>(64.1) | 0.96 (0.77-1.19),<br>p=0.692 | 701/910<br>(77.0) | 1.00 (0.76-1.31),<br>p=0.995 |
| **6-10 years** | 275/810<br>(34.0) | 0.85 (0.68-1.07),<br>p=0.172 | 269/810<br>(33.2) | 0.96 (0.77-1.21),<br>p=0.746 | 373/810<br>(46.0) | 1.06 (0.85-1.31),<br>p=0.606 | 509/810<br>(62.8) | 1.01 (0.80-1.27),<br>p=0.945 | 587/810<br>(72.5) | 1.05 (0.79-1.39),<br>p=0.732 |
| **>10 years** | 320/877<br>(36.5) | 0.78 (0.62-1.00),<br>p=0.047 | 308/877<br>(35.1) | 0.97 (0.77-1.23),<br>p=0.815 | 393/877<br>(44.8) | 0.93 (0.74-1.17),<br>p=0.538 | 543/877<br>(61.9) | 0.83 (0.65-1.05),<br>p=0.124 | 592/877<br>(67.5) | 0.94 (0.70-1.25),<br>p=0.656 |

Participants with no formal education reported the highest proportion of problems in mobility (48.3%), self-care (45.2%), and usual activities (54.8%). Conversely, participants with higher education had lower prevalence of difficulties, particularly in self-care (33.3%) and pain/discomfort (59.3%).

Employment status was strongly associated with HRQoL. Unemployed participants had markedly worse outcomes, reporting significantly higher rates of problems in self-care (43.9%), usual activities (57.9%), and anxiety/depression (80.9%). In contrast, employed individuals reported the lowest prevalence of problems across all domains, particularly in self-care (24.0%) and usual activities (32.0%).

Stratification by income tertile revealed further disparities. Participants in the lowest income group reported the highest proportions of impairment in all domains, particularly in mobility (37.6%), usual activities (48.1%), and anxiety/depression (77.0%). While those in the highest tertile still reported considerable levels of impairment, the figures were lower, especially for pain/discomfort (59.1%) and anxiety/depression (69.8%).

### Regression analysis model HRQoL with associated factors of HRQoL

Multivariable logistic regression models were used to examine the adjusted associations between sociodemographic, clinical, and behavioural variables and HRQoL problems across each EQ-5D-5L domain (Table 3). Compared to participants in Bangladesh, those in India had significantly lower odds of problems across all five domains, particularly for anxiety/depression (OR = 0.07, 95% CI = 0.05–0.09, p < 0.001) and pain/discomfort (OR = 0.30, 95% CI = 0.24–0.37, p < 0.001). Participants in Pakistan had significantly higher odds of reporting difficulties, especially with mobility (OR = 2.47, 95% CI = 1.98–3.08, p < 0.001) and usual activities (OR = 1.76, 95% CI = 1.42–2.18, p < 0.001).

Female participants had higher odds of experiencing pain/discomfort (OR = 1.51, 95% CI = 1.19–1.91, p < 0.001) and anxiety/depression (OR = 1.66, 95% CI = 1.24–2.23, p < 0.001) compared to males.

Older age groups showed progressively higher odds of HRQoL impairment. Participants aged 55+ had the highest odds of mobility issues (OR = 2.70, 95% CI = 1.89–3.85, p < 0.001) and pain/discomfort (OR = 2.10, 95% CI = 1.44–3.07, p < 0.001) compared to those aged 18–24.

Unemployed individuals had substantially higher odds of reporting HRQoL difficulties across all domains. For example, the odds of self-care problems were more than twice that of employed participants (OR = 2.30, 95% CI = 1.88–2.81, p < 0.001), and odds of anxiety/depression were also elevated (OR = 1.61, 95% CI = 1.27–2.05, p < 0.001).

High income was associated with beneficial effects across multiple domains. Compared to the low-income group, individuals in the high-income tertile had significantly lower odds of problems in pain/discomfort (OR = 0.65, 95% CI = 0.53–0.79, p < 0.001) and anxiety/depression (OR = 0.70, 95% CI = 0.56–0.88, p = 0.003). The middle-income group, however, had significantly higher odds of mobility (OR = 1.27, 95% CI = 1.06–1.52, p = 0.009) and self-care problems (OR = 1.21, 95% CI = 1.01–1.45, p = 0.038) compared to the low-income group.

Higher education levels were associated with significantly lower odds of HRQoL impairments. Participants with secondary education had reduced odds of problems in self-care (OR = 0.64, 95% CI = 0.48–0.84, p = 0.002), usual activities (OR = 0.65, 95% CI = 0.50–0.85, p = 0.002), and pain/discomfort (OR = 0.72, 95% CI = 0.54–0.97, p = 0.032), relative to those with no formal education.

Underweight participants had marginally higher odds of reporting mobility problems (OR = 1.30, 95% CI = 0.98–1.73, p = 0.072) compared to those with normal BMI. Interestingly, overweight individuals had significantly lower odds of self-care problems (OR = 0.78, 95% CI = 0.65–0.94, p = 0.009), suggesting a potential “obesity paradox” in this population.

Smokers had significantly higher odds of self-care (OR = 1.34, 95% CI = 1.13–1.59, p < 0.001) and usual activity problems (OR = 1.35, 95% CI = 1.15–1.59, p < 0.001). Participants with hypertension had increased odds of reporting pain/discomfort (OR = 1.30, 95% CI = 1.03– 1.65, p = 0.028) and anxiety/depression (OR = 1.30, 95% CI = 1.01–1.69, p = 0.042). High triglycerides were also associated with greater problems in usual activities (OR = 1.19, 95% CI = 1.02–1.38, p = 0.027). Inpatients reported higher impairment across all domains than outpatients. For example, inpatients had 43% higher odds of pain/discomfort (reference OR = 1.0; outpatient OR = 0.43, 95% CI = 0.34–0.54, p < 0.001).

### Visual Analogue Scale (VAS) Exclusion

While the EQ-5D-5L includes a Visual Analogue Scale (VAS) to capture participants’ self-rated health status, we excluded these data from the analysis. The VAS data in this study were affected by substantial variability, low correlation with EQ-5D domain scores, and site-level inconsistencies in administration and interpretation. Including VAS results risked compromising the clarity and comparability of the findings; therefore, we opted for a more conservative, domain-focused analytical approach.

## Discussion

This study provides insights into the prevalence of problems related to HRQoL and the factors associated with HRQoL among people with SMI in South Asia. The findings show substantial socioeconomic, demographic, and health-related disparities affecting HRQoL. Our results emphasise the role of income, education, employment, gender, multimorbidity, smoking, and body weight in shaping HRQoL outcomes and provide evidence that could inform practice, policy, and the development of targeted interventions.

### Socioeconomic Determinants of HRQoL

Our findings underscore the substantial influence of socioeconomic status, particularly income, employment, and education, on HRQoL among individuals with SMI. Participants in the highest income tertile were significantly less likely to report problems with usual activities, pain/discomfort, and anxiety/depression, suggesting that greater financial resources may help buffer against some of the adverse effects of SMI on quality of life. Likewise, individuals who were employed experienced fewer limitations across all HRQoL domains compared to their unemployed or economically inactive counterparts, highlighting the critical role of economic engagement in supporting functional well-being. Additionally, higher educational attainment appeared to be associated with improved outcomes, suggesting that education may confer resilience or better coping mechanisms in managing both physical and psychological health challenges.

These results align with previous studies from both LMICs and high-income countries (HICs), which consistently show that individuals with higher socioeconomic status are more likely to engage with health services, afford medications, and maintain better overall health [28,65,66]. Financial barriers are especially critical in LMICs, where out-of-pocket expenses and limited insurance coverage deter lower-income individuals from seeking psychiatric care, ultimately contributing to worse mental health outcomes [32,66].

Employment plays a dual role in supporting HRQoL. Beyond financial stability, it offers routine, social integration, and a sense of purpose—factors known to protect mental well-being. A systematic review by Modini et al. (2016) found that employment enhances mental health by reducing social isolation and improving self-esteem, particularly in individuals with psychiatric disorders [66]. Similarly, Bouwmans et al. (2015) reported that employed individuals with schizophrenia experienced greater life satisfaction and fewer depressive symptoms compared to their unemployed peers [29].

Moreover, individuals in higher-income groups are more likely to engage in health-promoting behaviours such as regular physical activity, nutritious diets, and the use of preventive health services, all of which contribute to better long-term health and quality of life [30,66]. Together, these findings reinforce the importance of addressing income and employment disparities as part of any comprehensive strategy to improve HRQoL among people with SMI.

### Geographic Variations in HRQoL

This study revealed significant regional disparities in HRQoL across India, Pakistan, and Bangladesh, likely reflecting differences in healthcare infrastructure, economic development, and mental health policy implementation. Notably, mental health treatment coverage is considerably higher in India (83.5%) compared to Pakistan (50.4%) and Bangladesh (44.8%) [31], suggesting that broader systemic investments in healthcare may contribute to better HRQoL outcomes.

India’s relatively stronger economic growth and expanding healthcare infrastructure appear to support higher HRQoL scores, particularly in domains such as mobility, self-care, and psychological well-being [45,70,72,73]. These findings align with research demonstrating better health outcomes among older adults and individuals with chronic illnesses in India, highlighting the important role of national context in shaping health experiences [45,70].

However, despite these regional differences, the gap in HRQoL between individuals with SMI and the general population remains substantial across all three countries. This suggests that serious mental illness exerts a profound and consistent burden on quality of life, regardless of national economic or healthcare conditions.

The observed trends also mirror disparities found in other vulnerable groups across South Asia, indicating that broader structural and policy-level interventions are needed to address the health and quality of life challenges faced by people with SMI in under-resourced settings [67,71].

### Gender Disparities in HRQoL

Gender emerged as a significant factor influencing HRQoL, with women more likely than men to report problems in the pain/discomfort and anxiety/depression domains, even after controlling for age, income, and comorbidities. This finding supports existing evidence from both LMICs and HICs, suggesting that women with SMI often experience a dual burden: increased psychiatric vulnerability compounded by social, cultural, and economic disadvantages [43,44,45,79].

In South Asia, women with SMI face multiple intersecting barriers, including stigma, limited autonomy, and restricted access to healthcare, all of which can intensify symptoms and lower quality of life [44,80]. Traditional gender roles in the region often place caregiving and domestic responsibilities disproportionately on women, leading to chronic stress and reduced opportunities for self-care, further diminishing their HRQoL [44,61].

While gender disparities in HRQoL exist globally, their manifestations differ across contexts. In LMICs, inadequate healthcare access and weaker social support systems amplify these disparities [43,44]. In contrast, in higher-income settings, improved gender equity, broader healthcare access, and stronger social protections help mitigate the negative impacts on women’s quality of life. European studies have shown that although women with depression or schizophrenia report lower HRQoL than men, the gender gap narrows substantially after accounting for healthcare access, financial independence, and employment status [73,81]. Similarly, research from North America highlights the role of workplace protections, mental health insurance, and gender-sensitive interventions in reducing disparities commonly observed in LMICs [74,82].

### Physical Health Conditions and HRQoL

Multimorbidity significantly impacted HRQoL, particularly in self-care and usual activities. These findings align with previous research indicating that chronic conditions exacerbate functional limitations, increase healthcare utilization, and significantly impair HRQoL [37,38]. The presence of multiple chronic illnesses, especially among populations with severe mental illness, further compounds the burden, reducing quality of life and complicating treatment outcomes [65,67]. Moreover, socioeconomic factors influencing access to care in low- and middle-income countries contribute to these adverse effects, underscoring the need for integrated healthcare approaches [63,65].

### Smoking and HRQoL

Smoking was strongly associated with lower HRQoL, particularly in self-care and usual activities. In this study, 34.5% of participants reported being smokers, substantially higher than the general South Asian population (12–25%) [75,77]. This aligns with global evidence showing elevated smoking rates among individuals with SMI, who are twice as likely to smoke as the general population [78–82]. High smoking prevalence contributes to increased risks of cardiovascular and respiratory diseases, multimorbidity, and premature mortality, all of which negatively impact quality of life [62,80,81]. Moreover, continued smoking has been linked to poorer psychiatric outcomes, including worsened depression, anxiety, and cognitive function [74,82]. These findings underscore the need for targeted smoking cessation interventions as part of comprehensive mental healthcare strategies in South Asia [79,83].

### The Role of BMI in HRQoL

This study adds important regional insight into the relationship between body weight and HRQoL among individuals with SMI in South Asia. Notably, underweight status was more strongly associated with impairments in mobility, self-care, and usual activities than overweight or obesity. These findings highlight the often-overlooked burden of undernutrition and frailty in LMICs, where food insecurity, poor physical health, and social isolation are prevalent among individuals with mental illness [33,34,35]. In people with SMI, being underweight may reflect a combination of factors such as malnutrition, adverse medication effects (e.g., appetite suppression), poor treatment adherence, and limited access to care, all of which may compromise physical and mental health. These vulnerabilities are often magnified by poverty and marginalisation, particularly in low-resource settings. The association between underweight status and poor HRQoL thus represents a complex interplay—often described as a “syndemic” of nutritional, psychological, and socioeconomic disadvantage [20,33,41].

Interestingly, overweight and obesity were not consistently linked to lower HRQoL in this sample. In some domains, such as self-care, being overweight appeared protective. This aligns with the so-called “obesity paradox,” where higher BMI may be associated with better health outcomes in certain contexts [36,38]. In LMICs, including South Asia, a higher BMI may indicate better socioeconomic status, greater food security, or access to healthcare—factors that can buffer against the negative health and psychosocial consequences typically associated with obesity in HICs [28,30,31,63,64,65]. Taken together, these findings reflect the “double burden of malnutrition”—the coexistence of underweight and overweight within the same population, which poses unique challenges for mental health and public health policy in many LMICs [28,34,58]. While overweight status may have some short-term associations with better functioning, the long-term cardiometabolic risks of obesity remain significant and warrant attention in integrated care approaches [62].

### Implications for Practice

This study highlights the urgent need for comprehensive, integrated strategies to improve HRQoL among individuals with SMI in South Asia. People with SMI who face socioeconomic disadvantage, physical comorbidities, and gender-based inequities are especially vulnerable and require tailored support across multiple domains of care.

In particular, women, individuals with chronic illnesses, and those from lower-income backgrounds experience compounded barriers to care and poorer HRQoL outcomes. Gender-sensitive interventions that address caregiving burdens, economic exclusion, and limited healthcare access are critical, especially in LMICs where cultural and structural barriers disproportionately impact women.

Expanding access to community-based mental health services and integrating psychiatric care into primary healthcare systems in Pakistan and Bangladesh is essential to improve early diagnosis, continuity of care, and access to psychosocial support. Strengthening these systems will reduce treatment gaps and support more equitable health outcomes across the region.

Employment and financial stability are key social determinants of mental health. Inclusive employment policies, vocational rehabilitation, and social protection programs, such as conditional cash transfers, could support recovery and social reintegration for individuals with SMI. These policies are particularly vital in regions where economic inequality continues to undermine both physical and mental well-being.

The high prevalence of physical health conditions such as diabetes, hypertension, and smoking-related illnesses among individuals with SMI underscores the need for holistic, integrated care models. Mental health services must routinely include screening and management for physical conditions, supported by collaborative care approaches and health system strengthening.

Targeted smoking cessation programs are also urgently needed, given the significantly elevated smoking rates among people with SMI. These interventions should be embedded within mental health services and designed to address both behavioral and structural drivers of tobacco use, with access to pharmacological and psychosocial support. Research should continue to explore the long-term benefits of smoking cessation on HRQoL in this population. Nutrition is another crucial but often neglected determinant of HRQoL. Interventions should be context-specific and nutritionally inclusive, ranging from meal support and micronutrient supplementation for underweight individuals to culturally appropriate dietary counseling and physical activity programs for those who are overweight or obese. All efforts should be situated within a broader psychosocial care model that addresses upstream factors such as poverty, stigma, and social exclusion.

Ultimately, a multi-sectoral approach that includes increased investment, system integration, and policy reform is needed to reduce HRQoL disparities. By identifying and targeting the most vulnerable subgroups, health systems in South Asia can better promote long-term well-being and functional recovery for individuals living with SMI.

## Limitations

This study is cross-sectional in nature, which limits the ability to infer causality. Additionally, reliance on self-reported data may introduce recall or social desirability bias. Country-specific contextual differences, such as the timing of data collection (pre- vs post-pandemic) and local service structures, may also influence the generalisability of the findings. Finally, the exclusion of VAS data, while necessary, may limit comparability with some prior EQ-5D studies.

## Conclusion

This study highlights significant and multifaceted disparities in HRQoL among individuals with SMI in South Asia. Key determinants include income, employment, gender, smoking, multimorbidity, and underweight status. These findings call for a multidimensional, equity-oriented approach to mental health care, one that integrates physical health management, promotes social inclusion, and addresses the broader determinants of well-being.

## Acknowledgments

The authors would like to acknowledge Abu Musa Robin, Anjuman Jum Tithi, Tanvir Arafat, Lipon Saha, Tasmia Rahman, Dr. Md. Helaluddin, Dr. Khaleda Islam, Dr. Mamun, and Dr. Bappi from the Bangladesh team; Archith Krishna, Sathish Kumar, Neeta P.S., Venkatalakshmi C., Bhuvneshwari L., Manjunatha S., Sobin George, and Krishna Jayanthi from the India team; and Rubab Ayesha, Nida Afsheen, Najma Hayat, Zaheen Amin, and Aniqa Maryam from the Pakistan team for conducting all interviews and physical measurements, as well as providing technical and managerial support. The authors also thank all the participants who consented and generously provided their time to complete the interviews.

## Author’s contributions

Conceptualisation: Avantika Sharma (AS). Methodology: Avantika Sharma (AS). Software: Alex Mitchell (AM), Fraser Wiggins (FW). Validation: Alex Mitchell (AM), Fraser Wiggins (FW). Formal analysis: Alex Mitchell (AM), Fraser Wiggins (FW). Investigation: Avantika Sharma (AS). Resources: Gerardo A Zavala (GAZ). Data curation: Krishna Prasad (KP), Kavindu Appuhamy (KA). Writing – original draft: Avantika Sharma (AS). Writing – review & editing: Badur Un Nisa (BN), Humaira Bibi (HB). Visualisation: Avantika Sharma (AS). Supervision: Gerardo A Zavala (GAZ). Project administration: Gerardo A Zavala (GAZ). Funding acquisition: Gerardo A Zavala (GAZ).

## Reference

1. Karimi M, Brazier J. Health, Health-Related Quality of Life, and Quality of Life: What is the difference? Pharmacoeconomics. 2016;34(7):645–9.

2. van de Willige G, Wiersma D, Nienhuis FJ, Jenner JA. Changes in quality of life in chronic psychiatric patients: a comparison between EuroQol (EQ-5D) and WHOQoL. Qual Life Res. 2005;14(2):441–51.

3. Folsom DP, Depp C, Palmer BW, et al. Physical and mental health-related quality of life among older people with schizophrenia. Schizophr Res. 2009;108(1–3):207–13.

4. Katz DA, McHorney CA, Atkinson RL. Impact of obesity on health-related quality of life in patients with chronic illness. J Gen Intern Med. 2000;15(11):789–96.

5. Kim SR, Kim HN, Song SW. Associations between mental health, quality of life, and obesity/metabolic risk phenotypes. Metab Syndr Relat Disord. 2020;18(7):347–52.

6. Singh K, Kaur J. India, quality of life. In: Michalos AC, editor. Encyclopedia of quality of life and well-being research. Dordrecht: Springer; 2014. p. 3187–90.

7. Ball K, Crawford D, Jeffery R, Brug J. The role of socio-cultural factors in the obesity epidemic. In: Crawford D, Jeffery RW, Ball K, Brug J, editors. Obesity epidemiology: from aetiology to public health. 2nd ed. Oxford: Oxford University Press; 2010. p. 105–18.

8. Hwalla N, Nasreddine L, El Labban S. Cultural determinants of obesity in low- and middle-income countries in the Eastern Mediterranean Region. In: Romieu I, Dossus L, Willett WC, editors. Energy balance and obesity. Lyon: International Agency for Research on Cancer; 2017. p. 1187. doi:10.26719/2012.18.12.1187

9. Rathod S, Pinninti N, Irfan M, et al. Mental health service provision in low- and middle-income countries. Health Serv Insights. 2017;10:1178632917694350.

10. Appuhamy KK, Podmore D, Mitchell A, et al. Risk factors associated with overweight and obesity in people with severe mental illness in South Asia: cross-sectional study in Bangladesh, India, and Pakistan. J Nutr Sci. 2023;12:e116.

11. Bartlem KM, Bowman JA, Bailey JM, et al. Chronic disease health risk behaviours amongst people with a mental illness. Aust N Z J Psychiatry. 2015;49(8):731–41.

12. Vallis M. Quality of life and psychological well-being in obesity management: improving the odds of success by managing distress. Int J Clin Pract. 2016;70(3):196– 205.

13. Rajan S, Mitchell A, Zavala GA, et al. Tobacco use in people with severe mental illness: findings from a multi-country survey of mental health institutions in South Asia. Tob Induc Dis. 2023;21:166.

14. Misra A, Tandon N, Ebrahim S, et al. Diabetes, cardiovascular disease, and chronic kidney disease in South Asia: current status and future directions. BMJ. 2017;357:j1420.

15. Umar Z, Tahir Z, Nizami A (2023) Impact of severe mental illnesses on health-related quality of life among patients attending the Institute of Psychiatry, Rawalpindi from 2019 to 2021: A cross-sectional study. PLOS ONE 18(8): e0289080. 10.1371/journal.pone.0289080

16. Zavala GA, Prasad-Muliyala K, Aslam F, et al. Prevalence of physical health conditions and health risk behaviours in people with severe mental illness in South Asia: protocol for a cross-sectional study (IMPACT SMI survey). BMJ Open. 2020;10(9):e037869.

17. Chand P, Murthy P, Arunachalam V, Naveen Kumar C, Isaac M. Service utilization in a tertiary psychiatric care setting in South India. Asian J Psychiatr. 2010;3(4):222–6.

18. World Health Organization. International statistical classification of diseases and related health problems 10th revision (ICD-10) version:2010. Available from: https://icd.who.int/browse10/2010/en [cited 2023 Jun 2].

19. Zavala GA, Todowede O, Mazumdar P, et al. Effectiveness of interventions to address obesity and health risk behaviours among people with severe mental illness in low- and middle-income countries (LMICs): a systematic review and meta-analysis. Global Mental Health. 2022;9:264–73.

20. Lecrubier Y, Sheehan DV, Weiller E, et al. The Mini International Neuropsychiatric Interview (MINI). A short diagnostic structured interview: reliability and validity according to the CIDI. Eur Psychiatry. 1997;12(5):224–31.

21. De Hert M, Schreurs V, Vancampfort D, Van Winkel R. Metabolic syndrome in people with schizophrenia: a review. World Psychiatry. 2009;8(1):15–22.

22. Buderer NM. Statistical methodology: I. Incorporating the prevalence of disease into the sample size calculation for sensitivity and specificity. Acad Emerg Med. 1996;3(9):895–900.

23. Bonita R, De Courten M, Dwyer T, Jamrozik K, Winkelmann R. Surveillance of risk factors for noncommunicable diseases: the WHO STEPwise approach: summary. Geneva: World Health Organization; 2001. Available from: https://digital.library.adelaide.edu.au/dspace/handle/2440/47457

24. Stephenson J, Smith CM, Kearns B, Haywood A, Bissell P. The association between obesity and quality of life: a retrospective analysis of a large-scale population-based cohort study. BMC Public Health. 2021;21(1):1990.

25. Deurenberg P, Deurenberg-Yap M, Guricci S. Asians are different from Caucasians and from each other in their body mass index/body fat per cent relationship. Obes Rev. 2002;3(3):141–6.

26. WHO Expert Consultation. Appropriate body-mass index for Asian populations and its implications for policy and intervention strategies. Lancet. 2004;363(9403):157– 63.

27. Pan WH, Flegal KM, Chang HY, Yeh WT, Yeh CJ, Lee WC. Body mass index and obesity-related metabolic disorders in Taiwanese and US whites and blacks: implications for definitions of overweight and obesity for Asians. Am J Clin Nutr. 2004;79(1):31–9.

28. Singh K, Kondal D, Shivashankar R, et al. Health-related quality of life variations by sociodemographic factors and chronic conditions in three metropolitan cities of South Asia: the CARRS study. BMJ Open. 2017;7(10):e018424. doi:10.1136/bmjopen-2017-018424

29. Bouwmans C, de Sonneville C, Mulder CL, Hakkaart-van Roijen L. Employment and the associated impact on quality of life in people diagnosed with schizophrenia. Neuropsychiatr Dis Treat. 2015;11:2125–42.

30. Ray TK, Kenigsberg TA, Pana-Cryan R. Employment arrangement, job stress, and health-related quality of life. Saf Sci. 2017;100:46–56.

31. Pinto AD, Hassen N, Craig-Neil A. Employment interventions in health settings: a systematic review and synthesis. Ann Fam Med. 2018;16(5):447–60.

32. Rahman MM, Karan A, Rahman MS, et al. Progress toward universal health coverage: a comparative analysis in 5 South Asian countries. JAMA Intern Med. 2017;177(9):1297–305.

33. Khan BA, Khalid H, Siddiqi N, et al. Prevalence of underweight in people with severe mental illness: systematic review and meta-analysis. Ment Health Sci. 2022; published online Dec 11. doi:10.1002/mhs2.7

34. Biswas T, Townsend N, Magalhaes R, Hasan MM, Mamun AA. Geographical and socioeconomic inequalities in the double burden of malnutrition among women in Southeast Asia: a population-based study. Lancet Reg Health Southeast Asia. 2022;1:100007.

35. Mistry SK, Puthussery S. Risk factors of overweight and obesity in childhood and adolescence in South Asian countries: a systematic review of the evidence. Public Health. 2015;129(2):200–9.

36. Bhurosy T, Jeewon R. Overweight and obesity epidemic in developing countries: a problem with diet, physical activity, or socioeconomic status? ScientificWorldJournal. 2014;2014:964236.

37. Makovski TT, Schmitz S, Zeegers MP, Stranges S, van den Akker M. Multimorbidity and quality of life: systematic literature review and meta-analysis. Ageing Res Rev. 2019;53:100903.

38. Afzal M, Siddiqi N, Ahmad B, et al. Prevalence of overweight and obesity in people with severe mental illness: systematic review and meta-analysis. Front Endocrinol (Lausanne). 2021;12:769309.

39. Faisal MR, Salam FT, Vidyasagaran AL, et al. Collaborative care for common mental disorders in low- and middle-income countries: a systematic review and meta-analysis. 2024 Jul 20 [published online]. Available from: https://scholar.google.com/citations?view_op=view_citation&hl=en&citation_for_view=xDU9-kQAAAAJ:blknAaTinKkC

40. Moly KT, Abraham D, Ashika MS. Quality of life in obese patients–gender differences. Indian J Public Health Res Dev. 2021;12(3):597–602.

41. Pimenta FB, Bertrand E, Mograbi DC, Shinohara H, Landeira-Fernandez J. The relationship between obesity and quality of life in Brazilian adults. Front Psychol. 2015;6:966.

42. Torres KD, Rosa ML, Moscavitch SD. Gender and obesity interaction in quality of life in adults assisted by a family doctor program in Niterói, Brazil. Cien Saude Colet. 2016;21(5):1617–24.

43. Almeida M, Sun JF. Serious mental illness in women. Curr Opin Psychiatry. 2022;35(3):157–64.

44. Ghebrehiwet S, Baul T, Restivo JL, Kelkile TS, Stevenson A, Gelaye B, et al. Gender-specific experiences of serious mental illness in rural Ethiopia: A qualitative study. Glob Public Health. 2020;15(2):185–99.

45. Lee KH, Xu H, Wu B. Gender differences in quality of life among community-dwelling older adults in low- and middle-income countries: Results from the Study on Global AGEing and Adult Health (SAGE). BMC Public Health. 2020;20(1):114.

46. Jyani G, Prinja S, Garg B, Kaur M, Grover S, Sharma A, et al. Health-related quality of life among Indian population: The EQ-5D population norms for India. J Glob Health. 2023;13:04018. doi:10.7189/jogh.13.04018.

47. Feng Y, Devlin N, Herdman M. Assessing the health of the general population in England: How do the three- and five-level versions of EQ-5D compare? Health Qual Life Outcomes. 2015;13:171. doi:10.1186/s12955-015-0356-8.

48. McCaffrey N, Kaambwa B, Currow DC, Ratcliffe J. Health-related quality of life measured using the EQ-5D-5L: South Australian population norms. Health Qual Life Outcomes. 2016;14:133. doi:10.1186/s12955-016-0537-0.

49. Yang Z, Busschbach J, Liu G, Luo N. EQ-5D-5L norms for the urban Chinese population in China. Health Qual Life Outcomes. 2018;16:210. doi:10.1186/s12955-018-1036-2.

50. Kim TH, Jo MW, Lee SI, Kim SH, Chung SM. Psychometric properties of the EQ-5D-5L in the general population of South Korea. Qual Life Res. 2013;22:2245–53. doi:10.1007/s11136-012-0331-3.

51. Hinz A, Kohlmann T, Stöbel-Richter Y, Zenger M, Brähler E. The quality of life questionnaire EQ-5D-5L: psychometric properties and normative values for the general German population. Qual Life Res. 2014;23(2):443–7. doi:10.1007/s11136-013-0498-2

52. Scalone L, Cortesi PA, Ciampichini R, Cesana G, Mantovani LG. Health-related quality of life norm data of the general population in Italy: Results using the EQ-5D-3L and EQ-5D-5L instruments. Epidemiol Biostat Public Health. 2015;12:e114571–1.

53. Yang Z, Busschbach J, Liu G, Luo N.EQ-5D-5L norms for the urban Chinese population in China. Health Qual Life Outcomes. 2018;16:210. 10.1186/s12955-018-1036-2.

54. Patnaik I, Sane R, Shah A, Subramaniam SV. Distribution of self-reported health in India: The role of income and geography. Plos one. 2023 Jan 27;18(1):e0279999.

55. Wan Puteh SE, Siwar C, Zaidi MAS, Kadir HA. Health related quality of life (HRQOL) among low socioeconomic population in Malaysia. BMC Public Health. 2019;19(Suppl 4):551.

56. National Institute of Mental Health (NIMH). Mental illness statistics [Internet]. 2023 [cited 2025 Jun 10]. Available from: https://www.nimh.nih.gov/health/statistics/mental-illness

57. World Health Organization (WHO). Mental disorders [Internet]. 2022 [cited 2025 Jun 10]. Available from: https://www.who.int/news-room/fact-sheets/detail/mental-disorders

58. Louie JC, Mills KT, Teasdale BJ, Kit BK. Double burden of malnutrition in Asia: Trends, policy responses, and the way forward. Lancet. 2009;374(9705):970–81.

59. Saxena S, Misra PJ, Vishwanath NS, Varma RP, Soman B. Quality of life and its correlates in Central India. Int J Res Dev Health. 2013;1(2):85–96.

60. Saha A, Muhammad T, Mandal B, Adhikary M, Barman P. Socio-demographic and behavioral correlates of excess weight and its health consequences among older adults in India: Evidence from a cross-sectional study, 2017–18. PLoS One. 2023;18(10):e0291920.

61. Kanter R, Caballero B. Global gender disparities in obesity: A review. Adv Nutr. 2012;3(4):491–8. doi:10.3945/an.112.002063.PMID: 22797984

62. Specchia ML, Veneziano MA, Cadeddu C, Ferriero AM, Mancuso A, Ianuale C, et al. Economic impact of adult obesity on health systems: A systematic review. Eur J Public Health. 2015;25:255–62.

63. Dinsa GD, Goryakin Y, Fumagalli E, Suhrcke M. Obesity and socioeconomic status in developing countries: A systematic review. Obes Rev. 2012;13(11):1067–79. doi:10.1111/j.1467-789X.2012.01017.x.

64. Popkin BM. The nutrition transition and obesity in the developing world. J Nutr. 2006;136(1):1–3.

65. Lund C, Breen A, Flisher AJ, Kakuma R, Corrigall J, Joska JA, et al. Poverty and common mental disorders in low- and middle-income countries: A systematic review. Soc Sci Med. 2010;71(3):517–28.

66. Patel V, Saxena S, Lund C, Thornicroft G, Baingana F, Bolton P, et al. The Lancet Commission on global mental health and sustainable development. Lancet. 2018;392(10157):1553–98.

67. Modini M, Joyce S, Mykletun A, Christensen H, Bryant RA, Mitchell PB, et al. The mental health benefits of employment: Results of a systematic meta-review. Aust Psychiatry. 2016;24(4):331–6.

68. Marmot M, Bell R. Fair society, healthy lives. Public Health. 2012;126(Suppl 1):S4–10.

69. Khan MM. The dire state of mental health services in Pakistan. Lancet Psychiatry. 2016;3(9):794–5.

70. Jyani G, Prinja S, Garg B, Kaur M, Grover S, Sharma A, et al. Health-related quality of life among Indian population: the EQ-5D population norms for India. J Glob Health. 2023;13:04018.

71. Siddiqui S, Khalid R, Javed N, et al. Health-related quality of life in Pakistani adults: Normative values using EQ-5D-5L. Health Qual Life Outcomes. 2021;19:162.

72. Saxena S, Misra PJ, Vishwanath NS, Varma RP, Soman B. Quality of life and its correlates in Central India. Int J Res Dev Health. 2013;1(2):85–96.

73. Van der Meer M, Tullius JM, Bouwmans C, et al. Gender differences in health-related quality of life in patients with mental illness: A European perspective. Soc Psychiatry Psychiatr Epidemiol. 2022;57(3):563–75.

74. Kessler RC, McLaughlin KA, Green JG, et al. Gender and depression: Epidemiology and treatment. Annu Rev Clin Psychol. 2010;6:139–68.

75. Sinha DN, Palipudi KM, Rolle I, Asma S. Tobacco use among youth and adults in member countries of the South-East Asia region: Review of findings from surveys under the Global Tobacco Surveillance System. Indian J Public Health. 2011;55(3):169–76.

76. Szatkowski L, McNeill A. Diverging trends in smoking behaviors according to mental health status. Nicotine Tob Res. 2015;17(3):356–60.

77. Olfson M, Gerhard T, Huang C, Crystal S, Stroup TS. Premature mortality among adults with schizophrenia in the United States. JAMA Psychiatry. 2015;72(12):1172– 81.

78. McClave AK, McKnight-Eily LR, Davis SP, Dube SR. Smoking characteristics of adults with selected lifetime mental illnesses: Results from the 2007 National Health Interview Survey. Am J Public Health. 2010;100(12):2464–72.

79. Tsoi DT, Porwal M, Webster AC. Interventions for smoking cessation and reduction in individuals with schizophrenia. Cochrane Database Syst Rev. 2010;(6):CD007253.

80. Lawrence D, Mitrou F, Zubrick SR. Smoking and mental illness: Results from population surveys in Australia and the United States. BMC Public Health. 2009;9:285.

81. Chesney E, Goodwin GM, Fazel S. Risks of all-cause and suicide mortality in mental disorders: A meta-review. World Psychiatry. 2014;13(2):153–60.

82. Taylor GMJ, Itani T, Thomas KH, Rai D, Jones T, Windmeijer F, et al. Prescribed medications and risk of relapse in schizophrenia: A population-based cohort study. BMJ Open. 2021;11(5):e045465.

83. Banham L, Gilbody S. Smoking cessation in severe mental illness: What works? Addiction. 2010;105(7):1176–89.

